# Structural determinants of Sexually Transmitted Infections (STI) service delivery for internally displaced persons in Nigeria: A qualitative study of policy and humanitarian perspectives

**DOI:** 10.64898/2026.03.16.26348567

**Authors:** Oluwakemi Amodu, Craig Janes, Precious Affia

**Affiliations:** Faculty of Nursing, University of Alberta, Edmonton, Alberta, Canada; University of Waterloo, Waterloo, Ontario, Canada; University of Alberta, Edmonton, Alberta, Canada

## Abstract

Sexually transmitted infections (STIs) remain a major global public health concern, with a disproportionate burden in low- and middle-income countries affected by conflict and displacement. In Nigeria, STI prevention and treatment sit within a crowded sexual and reproductive health (SRH) policy landscape shaped by vertically financed HIV programmes, expanding self-care agendas and one of the largest internally displaced populations in Africa. This qualitative study examines how structural, institutional and sociocultural forces shape STI service delivery for IDPs and how they reproduce or challenge sexual and reproductive health and rights. We conducted in-depth interviews with federal and state policymakers, United Nations representatives and national and international non-governmental actors involved in SRH and humanitarian programming in Nigeria. Using an interpretive, structurally informed approach, we explored participants’ accounts of funding architectures, governance priorities, humanitarian protocols, provider practices and gendered norms. Nigeria was considered “policy-rich but implementation-poor”, with HIV-centred vertical financing creating a hierarchy of infections that renders non-HIV STIs comparatively invisible, unsubsidised and often effectively privatised in displacement settings. Chronic commodity shortages, workforce depletion in conflict-affected areas, weak surveillance systems and reliance on informal providers were seen as routine features of STI care in IDP camps. Sociocultural dynamics, including toilet infection narratives, gendered gatekeeping of women’s healthcare and STI-related stigma, shaped how IDPs interpreted symptoms, when they sought care and which providers they used. At programme and government levels, self-care and task-shifting policies, although framed as expanding choice and autonomy, were implemented through fragile supply chains, limited regulation of informal providers and uneven access to digital platforms. The findings indicate that improving STI care for IDPs in Nigeria will require rebalancing HIV-dominated financing, securing affordable STI commodities, strengthening supervision and data systems and referral pathways so that self-care and humanitarian responses translate into accessible, reliable services.

## Introduction

Sexually transmitted infections (STIs) remain a major global public health challenge, with substantial morbidity concentrated in low- and middle-income countries. The World Health Organization estimates that in 2020 there were approximately 374 million new infections with one of four curable STIs, namely chlamydia, gonorrhoea, syphilis and trichomoniasis, among people aged 15 to 49 years [1]. Beyond acute clinical symptoms, untreated STIs are associated with serious long-term consequences including pelvic inflammatory disease, infertility, ectopic pregnancy, adverse pregnancy outcomes and increased susceptibility to HIV infection [2,1].

In response to persistent access barriers, the World Health Organization has increasingly promoted self-care interventions as a strategy to expand the reach of sexual and reproductive health (SRH) services, particularly in settings where health systems face structural constraints [3]. WHO guidance highlights interventions such as HIV self-testing, human papillomavirus (HPV) self-sampling and selected STI self-collection approaches as evidence-based tools that can improve autonomy and extend service coverage. However, these interventions are intended to complement functional health systems and require regulatory oversight, commodity security and effective referral pathways to ensure safe implementation [3].

Nigeria carries a substantial share of the STI and HIV burden in sub-Saharan Africa. Although nationally representative data on non-HIV STIs remain limited, available reports suggest high prevalence across both key populations and segments of the general population [4,1]. National HIV prevalence has declined to an estimated 1.3 to 2.1 percent among adults aged 15 to 49 years, yet the absolute number of people living with HIV remains among the highest globally, with women and young people disproportionately affected [5]. Facility-based studies and targeted surveillance continue to document concurrent infections and persistent STI risk across diverse population groups, suggesting ongoing transmission and under-recognised burden.

At the same time, Nigeria faces one of the largest internal displacement crises in Africa. As of 2024, estimates indicate more than three million internally displaced persons across the country, largely driven by insurgency, communal violence, banditry and environmental disasters [6]. Many displaced populations reside in overcrowded camps or informal settlements characterised by limited water and sanitation infrastructure, constrained access to healthcare services and heightened exposure to sexual and gender-based violence. Disrupted livelihoods, transactional sex, weakened social protection systems and barriers to healthcare access collectively create conditions that increase vulnerability to STIs and other SRH risks in displacement settings.

In response to these intersecting challenges, Nigeria has developed an extensive architecture of SRH policies and guidelines, including national HIV and STI strategies, task-shifting and task-sharing frameworks, and humanitarian commitments aligned with the Minimum Initial Service Package for reproductive health in emergencies [7,8]. More recently, the Federal Ministry of Health adopted the National Guideline on Self-Care for Sexual, Reproductive and Maternal Health for 2021 to 2026, aligning national policy with WHO recommendations and formally integrating self-care approaches into SRH programming [9,3]. These guidelines endorse interventions such as HIV self-testing and expanded community-level service delivery through pharmacies and patent medicine vendors.

Despite these policy developments, evidence suggests that STI services in Nigeria remain heavily HIV-centred, urban-biased and insufficiently integrated into humanitarian health responses. Surveillance systems for non-HIV STIs remain weak, national prevalence estimates are limited and little is known about how displaced populations access, or fail to access, STI prevention, testing and treatment [1]. Persistent gaps in commodities, workforce capacity and data systems are particularly pronounced in conflict-affected states, where health facilities are overstretched and reliance on lower-cadre providers and informal service channels is high.

These structural conditions raise important questions about how Nigeria’s evolving SRH policy landscape translates into practical access to STI care for internally displaced populations. While policy frameworks emphasise self-care, integration and community-based service delivery, their operational implications in humanitarian contexts remain insufficiently understood. The urgency of these questions has been sharpened by the suspension and termination of PEPFAR-funded programme activities in Nigeria following the United States foreign assistance freeze of January 2025, which abruptly disrupted antiretroviral supply chains and key population services across multiple states [10]. This development illustrates, in real time, the systemic fragility that vertically financed, donor-dependent HIV programming creates — and the compounding risk for displaced populations who have no alternative access pathway. A study that analyses the structural conditions underpinning this fragility, grounded in pre-disruption policy and programme perspectives, therefore speaks directly to present-day concerns about how Nigeria rebuilds and diversifies its SRH financing architecture.

This study addresses this gap by examining how national STI and SRH policies, including self-care initiatives, task-shifting strategies and humanitarian coordination mechanisms, shape STI service delivery for internally displaced persons in Nigeria. By foregrounding the perspectives of national policymakers, United Nations agencies and non-governmental organisations involved in SRH and humanitarian health programming, the study analyses the structural, policy and sociocultural determinants influencing STI prevention and care in displacement settings.

## Materials and Methods

### Study Design and settings

This study forms part of a broader qualitative inquiry examining sexual and reproductive health service delivery in Nigeria. The present analysis focuses specifically on how structural, institutional, and sociocultural determinants shape sexually transmitted infection (STI) service delivery for internally displaced persons (IDPs).

A qualitative interpretive approach was adopted to examine how policymakers, programme leaders, and humanitarian actors describe the organisation, financing, and implementation of STI and broader SRH services. The study was designed to capture institutional perspectives rather than individual patient experiences, with particular attention to how actors positioned within health governance structures interpret system constraints, funding hierarchies, and policy translation processes.

The analytic orientation draws on interpretive public health traditions and structural perspectives that situate health service delivery within broader political and economic systems [13,14]. This framework informed interpretation of how STI access is shaped by financing arrangements, governance priorities, and sociocultural dynamics.

Internal displacement is concentrated primarily in the North-East, North-West, and North-Central regions, where insecurity and conflict have disrupted health infrastructure and humanitarian service delivery [5,6].

STI service delivery operates within a mixed public–private health system characterised by vertically financed HIV programming, task-shifting and task-sharing policies, reliance on Patent and Proprietary Medicine Vendors (PPMVs), and the recent expansion of self-care strategies [15,9]. In displacement settings, services are delivered through government primary healthcare facilities, humanitarian agencies operating under cluster coordination mechanisms, and informal private providers. These intersecting systems formed the contextual backdrop of the study.

### Participants

The study draws on interviews with individuals occupying leadership and coordination roles within national and subnational STI, HIV, sexual and reproductive health (SRH), and humanitarian programming structures. Participants included federal-level officials within HIV/STI and SRH units, state primary healthcare leaders, representatives of United Nations agencies, international and national non-governmental organisations, and programme managers engaged in reproductive health coordination in displacement contexts. We conducted 11 semi-structured interviews with stakeholders involved in STI and broader secual and reproductive health (SRHR) policy and programming in Nigeria. Participants included two senior officials from the Federal Ministry of Health (one previously leading the national sexual and reproductive health portfolio, and one managing HIV/STI programming within the HIV/AIDS division), and one director from a Federal Capital Territory primary health care board responsible for overseeing primary-level services. Three participants were programme leads from Nigerian non-governmental organisations delivering sexual and reproductive health and humanitarian services, including projects in internally displaced persons’ camps funded by agencies such as United Nations Population Fund (UNFPA) and bilateral donors. Two participants were senior staff from international technical and implementing organisations working in Nigeria, with responsibilities for self-care policy, family planning and commodity security, and coordination of sexual and reproductive health interventions in humanitarian settings. The remaining three participants were practising sexual and reproductive health clinicians and clinical advisers (including obstetrician-gynaecologists) based in tertiary hospitals and primary health care facilities, with direct responsibility for maternal health, STI management and task-shifting supervision. For confidentiality, we report participants’ sectors and roles without naming individual facilities or specific programmes.

Recruitment for this project was conducted between 10/01/2022 and 15/03/2022. Data were collected through semi-structured, one-to-one interviews held virtually via telephone or videoconference between May and September 2022, a mode chosen to accommodate COVID-era travel constraints, the geographic dispersion of participants across Nigeria, and institutional preferences for remote engagement. All participants provided written consent by signing a consent form prior to their interview. Verbal consent was additionally obtained from each participant immediately before the commencement of their interview and documented by the interviewer as part of the study record. No independent witness was present during the verbal consent process. This consent procedure was approved by the University of Waterloo Research Ethics Board (Protocol #43034) and the National Health Research Ethics Committee of Nigeria (NHREC/01/01/2007).

### Data Analysis

Interview material was reviewed and analysed to identify recurring patterns in how participants described funding architecture, policy implementation, health system constraints, sociocultural dynamics, and emerging service delivery strategies. The analysis focused on institutional narratives and structural explanations rather than individual-level accounts. Through iterative engagement with the interview material, thematic patterns were identified across participants’ descriptions of STI programming in displacement settings. Particular attention was paid to how HIV and non-HIV STIs were positioned within financing and governance structures, how service delivery constraints were characterised, and how innovations such as self-care and digitalisation were framed. Interpretation was informed by scholarship on structural determinants of health and global health priority-setting framing, allowing the findings to be situated within broader debates on financing power, institutional visibility, and policy prioritisation [13,16].

### Conceptual Framework: Structural Determinants of STI Service Delivery

This study is grounded in a structural perspective on health, recognising that access to STI prevention and treatment is shaped less by individual behaviour alone and more by the systems within which people live. For internally displaced persons (IDPs) in Nigeria, STI service delivery unfolds within overlapping systems of health financing, governance arrangements, sociocultural norms, and humanitarian response mechanisms. Examining these interconnected layers helps explain why certain infections attract sustained institutional support while others remain marginal within policy and programming.

The analysis draws on the structural determinants of health framework as defined by Heller and colleagues [13], which conceptualises structural determinants as the written and unwritten rules—laws, policies, regulations, and deeply embedded norms—that create, maintain, or eliminate durable and hierarchical patterns of advantage among socially constructed groups in the conditions that affect health. Unlike earlier social determinants frameworks that risked reducing structure to decontextualised living conditions, this formulation centres power explicitly: structural arrangements are the products of political and economic agents whose decisions distribute visibility, resources, and protection unevenly across populations. In this study, the framework provides a lens for understanding how health financing architectures, governance priorities, legal constraints, and conflict-related instability produce patterned disadvantage in STI service access for internally displaced persons. Gaps in services are not merely administrative shortcomings; they reflect institutional logics, enacted through funding rules and procurement systems, that render certain infections politically durable and others structurally invisible.

The study also draws on recent scholarship on global health priority-setting framing [14,17], which demonstrates that diseases achieve political traction through three interlocking processes: securitisation (framing as an existential threat), moralisation (framing as an ethical imperative), and technification (framing as a cost-effective, scientifically solvable investment). Gómez and colleagues [14] show that these framing processes are not merely rhetorical; they mobilise elite coalitions within philanthropic foundations, international organisations, and donor agencies, which in turn shape which diseases become institutionally legible and which are sidelined. Complementing this, Smith and colleagues [17] demonstrate empirically that disease priority levels across agenda-setting arenas between 2000 and 2022 aligned neither with mortality burden nor with international norms alone, but reflected the relative organisational strength of advocacy networks and the durability of dedicated financing instruments. HIV’s institutional prominence in Nigeria exemplifies both dynamics. Over time, HIV programming secured dedicated funding streams, structured reporting requirements, and vertically organised supply chains, rendering it measurable, monitorable, and politically durable. Non-HIV STIs lack comparable coalitions and institutional safeguards, which affects the consistency with which they are financed, monitored, and integrated into routine and humanitarian services.

These theoretical perspectives informed the interpretation of participant narratives. When respondents described the fiscal privileging of HIV, the limited public procurement of non-HIV STI commodities, or the absence of reliable surveillance data in displacement settings, these accounts were examined as expressions of structural arrangements rather than isolated implementation failures. Similarly, discussions of stigma, “toilet infection” narratives, and male gatekeeping of women’s healthcare were understood within broader sociocultural systems that regulate sexuality and shape care-seeking practices. Humanitarian constraints, including insecurity, workforce depletion, and emergency-driven programming, were treated as additional structural layers that influence how national policies are enacted in IDP camps.

By situating the findings within these intersecting frameworks, the analysis moves beyond description to explanation. STI service delivery for displaced populations in Nigeria emerges as the outcome of interconnected financial, institutional, and social systems that collectively determine which health issues are prioritised, subsidised, and monitored, and which remain peripheral within the health architecture.

## Results

Analysis of the interviews revealed five interrelated structural dynamics shaping STI service delivery for internally displaced persons (IDPs) in Nigeria. These were: the structural dominance of HIV within national financing architectures; the disjuncture between policy commitments and implementation in IDP settings; persistent health system constraints related to commodities, workforce and data; sociocultural norms and gendered power relations that shape STI interpretation and care-seeking; and emerging but unevenly integrated innovations such as self-care and digital platforms. Across accounts, participants portrayed STI service delivery in displacement settings as governed less by written guidelines than by funding flows, institutional hierarchies and humanitarian logics, and the social conditions in which programmes operate.

### Funding architecture and the structural dominance of HIV

Participants consistently described a financing environment in which HIV programming occupies a structurally dominant position within Nigeria’s health system. While STIs are formally subsumed under broader sexual and reproductive health (SRH) programmes, donor earmarking, and vertically organised disease initiatives were said to create an implicit hierarchy of infections. A federal official summarised this dynamic: “By and large HIV has more support than STI program in Nigeria.” (Federal Ministry of Health – HIV/STI unit).

Respondents linked HIV’s dominance to long-standing international financing coalitions—such as PEPFAR and the Global Fund—that provide commodities, infrastructure and stable procurement systems, but also confer political visibility and administrative weight. As a senior staff member from a US-funded SRH implementing partner explained: “You would have organizations who are very interested in HIV/AIDS… significant funding from the Global Fund [and] the government of US… HIV has a lot of foot soldiers. Most STI are neglected diseases, let me put it like that.”

Several participants emphasised how vertical funding shapes internal government dynamics, with programmes operating in parallel silos rather than as an integrated system. One respondent commented that “the HIV/AIDS guys don’t want to hear anything outside of HIV, the guys on malaria don’t want to hear anything outside of malaria… what that does is that the other interventions are neglected… the STI unit is really overshadowed and government doesn’t quite invest in them.” (Senior staff, US-funded SRH implementing partner) At sub-national level, officials reported no dedicated budget lines for STIs outside HIV, and described STI care as largely dependent on general primary health care budgets and external projects. A primary health care director observed that “strictly about the line of funding specifically for STD/STIs apart from HIV I don’t think so.” (State Primary Health Care Board)

The practical consequence of this architecture was a bifurcated model of care. HIV treatment was widely described as free at the point of use, supported by donor-backed procurement and national supply chains, whereas treatment for other STIs typically required out-of-pocket payment for antibiotics and diagnostics. “For other STI generally they have to pay, they have to buy the antibiotics and they need to pay for that, it’s not free in most places,” noted a policy-level staff member at an international SRH NGO. Domestic mechanisms such as the Basic Health Care Provision Fund (BHCPF) were not perceived as capable of closing this gap in the short term; one respondent cautioned that expectations of the BHCPF as a new funding source were “overrated”, given how funds flow through insurance gateways and facility-level decision-making.

Across interviews, funding architecture was framed not only as a question of scarcity but as a determinant of visibility and rights. Conditions backed by vertical streams were counted, monitored and subsidised through dashboards and performance frameworks, while non-HIV STIs were more likely to remain invisible in national indicators and budget debates. In IDP settings, this translated into a situation where HIV-related care was publicly financed, and other STIs were effectively privatised, accessed through patent and proprietary medicine vendors (PPMVs) and private pharmacies on a fee-for-service basis.

### Policy abundance and implementation constraints in IDP settings

Participants repeatedly described Nigeria as a country with strong policy frameworks but persistently weak implementation. Federal officials and international partners highlighted an extensive SRH policy architecture, including national strategies, guidelines, protocols and training manuals that explicitly reference “vulnerable populations” and people in humanitarian settings. One international specialist remarked: “As a country we do have… the policy in place that support service delivery… you will find that the country doesn’t have issues with [policy], you will also find that the country has development partners that are supporting it.” (Senior staff, UN reproductive health agency). A Senior technical officer, WHO Nigeria (SRH and self-care policy) identified that “most of our SRH policy tools, strategic plans… have the vulnerable population including those in the IDP camp… the reproductive health service protocols and training manuals also include areas dealing with people that are internally displaced or generally people in humanitarian settings.”

Yet respondents drew a sharp distinction between this formal inclusion and operational delivery in camps. At implementation level, HIV services in some IDP settings were described as relatively well established, integrated into primary care and offered free of charge, while broader STI services were patchier and more dependent on local projects and commodity availability. As the HIV/STI Ministry official noted, “in IDP camps we still have some HIV services being offered in [an] integrated manner… the treatment is free and the drugs are still free… For STI as well it’s still the same process… but again the STI drugs [are] not as free as compared to HIV.” A primary health care director similarly noted that while HIV attracts substantial government and international investment — “money has been pumped by the government and international organization to tackle the issue of HIV” — dedicated funding lines for non-HIV STIs at the facility level were absent: “strictly about the line of funding specifically for STD/STIs apart from HIV, I don’t think so.” (Director, State Primary Health Care Board)

Camp-level shortages of medicines and diagnostics were common in participants’ accounts, driving reliance on PPMVs as frontline providers. “In the IDP camp the clinic does not really have enough medicine and they usually rely on the patent medicine vendors,” explained the primary health care director. Humanitarian coordination mechanisms—such as the Minimum Initial Service Package (MISP) for reproductive health—were recognised as conceptually comprehensive, but respondents stressed that emergency conditions, security concerns and workforce gaps limited the depth and consistency of implementation. A UN reproductive health agency representative described the MISP as “a holistic approach to deal with sexual reproductive health… in the case of STI, HIV kit 1 to 5,” yet emphasised that “you will find… limited health workers… many have been displaced… so you have to use simple tools and methodology like syndromic management which is not the gold standard but you have to use that because of the human resources available.”

Former federal officials also described fragmented governance for IDPs’ health, with responsibilities split between the Ministry of Humanitarian Affairs, the national emergency agency and the health ministry. One noted that emergency agencies prioritise shelter, water and security, and “when they remember health needs SRH will come very last,” leaving SRH in camps largely dependent on NGOs and private partners.

Overall, participants portrayed a narrowing process in which expansive national commitments are progressively filtered by financing, logistics and humanitarian priorities, resulting in a much thinner layer of STI services in IDP camps than policy language might suggest.

### Health system constraints: commodities, workforce and data Commodity insecurity and clinical spectrum of STIs

Across interviews, chronic shortages of STI diagnostics and antibiotics were described as routine, particularly in conflict-affected northern states. International and national actors pointed to stock-outs of test kits and treatment courses, exacerbated by insecurity, weak last-mile logistics and limited domestic financing. A state-level primary health care leader reported that “even the commodities, consumables and essential drugs are [in] shortage in our facilities.” (Director, State Primary Health Care Board) Respondents contrasted this with HIV, where antiretroviral therapy is largely donor-subsidised and channelled through robust HIV supply chains, while STI commodities must often be purchased by clients or procured ad hoc by facilities.

When asked which infections they were most concerned about, a national HIV/STI lead described STIs as “a broad range of disease… not only HIV, syphilis, gonorrhea, we have so many of them… also hepatitis B is a STD.” A tertiary gynaecologist characterised HPV as “the commonest STI” in her practice, linking it to high burdens of premalignant lesions and invasive cervical cancer among women from northern Nigeria, including those referred from IDP-hosting areas. She described “the regular diseases of poverty”—advanced cervical cancer, vulval disease and chronic pelvic pain—among women who often arrived late after repeated informal treatment.

Because culture and sensitivity testing for organisms such as Neisseria gonorrhoeae and Chlamydia trachomatis was rarely available, clinicians described relying on syndromic management and broad-spectrum “triple antibiotics” to “cover” likely chlamydial and gonococcal infections when women presented with heavy, malodorous discharge and lower abdominal pain. The gynaecologist stressed that “the laboratory does not have the facilities” and that the main problem was lack of diagnostics and culture media, rather than observed resistance to specific drugs. Some interviewees nonetheless worried that repeated empiric treatment of recurrent symptoms without laboratory confirmation could have implications for antimicrobial resistance, even though resistance testing for gonorrhoea was not routinely accessible.

### Workforce shortages and task-shifting

Participants highlighted the cumulative impact of conflict-related displacement, migration and “brain drain” on the health workforce. “When you have conflict-related crisis the human resources in those locations tend to migrate away… some of them have been virtually killed,” noted a senior staff member from a US-funded SRH implementing partner. In IDP-hosting areas, primary care facilities were described as relying heavily on community health extension workers (CHEWs) and, informally, PPMVs. The FCT primary health care director explained that “human resources for health generally in the entire nation is short… we use community health extension workers to manage the place,” particularly in remote communities where doctors and nurses are reluctant to be posted.

While task-shifting policies were viewed as necessary to maintain coverage, concerns were raised about the training, regulation and supervision of lower-cadre providers. A policy-level staff member at an international SRH NGO commented that people in rural communities “often depend on these PPMVs… the regulatory environment is the major challenge… another is the quality of services—some of them literally have primary school or secondary school education.” Federal respondents noted that recent revisions to task-shifting policy and PPMV regulation have sought to stratify PPMVs by training level and define their scope of practice, but acknowledged that enforcement and supervision remain uneven, especially in hard-to-reach areas.

### Data invisibility

Weaknesses in routine data systems for non-HIV STIs emerged as a further structural constraint. Participants observed that many STI episodes are managed in private pharmacies, PPMV shops or NGO clinics whose data may not feed into national health information systems. “Patent medicine vendors are the ones doing the assessment and providing the medication. So it’s very difficult… to say where the data are,” noted one international SRH policy specialist. A former federal SRH director described efforts to digitise logistics and service data for family planning, but noted that system expansion to include private providers has been technically challenging and remains incomplete. He argued that “if you really must encourage transmission of data, they should be able to transmit data right from the point of collection instead of going through all this paper-based collection… that can take like six months before the data will get to the federal level.”

Participants suggested that these commodity and workforce gaps are compounded by weak, largely paper-based information systems that exclude PPMVs and many humanitarian actors, reinforcing the statistical invisibility of non-HIV STIs in IDP settings and, in turn, their low political and budgetary priority.

### Sociocultural logics, gendered power and HPV vaccination

In addition to structural constraints, respondents described sociocultural narratives and gendered power dynamics that shape how STIs are understood and addressed in IDP contexts. A recurring theme was the attribution of genital symptoms to “toilet infection” or poor sanitation rather than sexual transmission, a framing that providers associated with limited health literacy but also with efforts to avoid sexual stigma. One international respondent remarked that “there really [is] no direct link between environmental sanitation and STI… I will simply limit it to ignorance,” though others suggested that these narratives can be understood as socially safer ways to discuss sexual health.

Clinicians working in and around camps described seeing large numbers of women with itching, discharge and pelvic pain who linked their symptoms to dirty toilets or poor personal hygiene. A public health physician explained that many women in her previous IDP camp study “had a lot of complain about itching, pain, discharge” and often attributed this to “bad toilet” rather than to sexual risks, even when they reported partner concurrency or sexual violence. These symptom narratives were seen as contributing to delayed care-seeking, repeated informal treatment and progression to complications such as infertility and cervical cancer.

Gendered power relations were central to accounts of access and decision-making. In some northern settings, women’s ability to seek screening or treatment was said to depend on male permission. A tertiary-level gynaecologist noted that “in the North a woman cannot come for screening if her husband does not permit her… male involvement is very key.” Participants also linked displacement with heightened exposure to transactional sex and gender-based violence, noting that economic precarity and insecurity can push women and adolescent girls into relationships that increase STI risk while limiting their bargaining power over condom use. “A lot happens around STI and gender-based violence around women who have transactional sex… when an STI is not treated on time… some people will still be living with that with infertility,” observed a policy-level NGO respondent.

HPV vaccination emerged as a further axis of inequity. A gynaecologist involved in national cervical cancer advocacy described strong demand for HPV vaccination among caregivers, but noted that, until very recently, vaccines were largely unavailable or prohibitively expensive, even for middle-class families. She recounted her own difficulty obtaining doses for her daughter and emphasised that girls in IDP camps were unlikely to access private vaccination. Although a national school-based HPV vaccination programme is planned, participants expressed concern that girls who are out of school, displaced or living in informal settlements could be bypassed unless specific outreach strategies are developed.

These accounts underscore how stigma, gendered authority structures, economic vulnerability and gaps in preventive technologies intersect with structural constraints to influence when and where displaced people seek care, which providers they trust, and which infections remain untreated or unprevented.

### Emerging innovations: self-care, telemedicine and digitalisation

Participants discussed emerging innovations—particularly self-care, telemedicine and digital tools—as potential avenues for expanding STI-related services, while cautioning that these remain unevenly integrated and largely focused on family planning and HIV. Self-care interventions, including contraceptive self-injection and HIV self-testing, were described as the “flagship” area of current activity. A self-care specialist from an international technical agency noted that “the flagship self-care intervention is more in the area of family planning, not treatment of STI,” even though STI self-testing was included in national guidance in a limited way.

Several interviewees described rapid expansion of digital health platforms during the COVID-19 period, including services that allow clients to consult clinicians by phone or online and receive recommended medications by delivery. One policy-level respondent explained that “there were some of these digital health platforms where people can chat with the doctor… they recommend and ship the right medication.” These platforms were viewed as particularly promising for urban populations and younger users, but respondents expressed uncertainty about their reach and appropriateness in camps and rural areas where literacy, connectivity and device access are limited. “I don’t know how well that is being used in IDP [settings], especially when you have people without formal education,” one interviewee cautioned.

Across interviews, digital and self-care innovations were framed as helpful but insufficient in isolation. Participants emphasised that, without stable commodity supply, regulation of private and community-level providers, integration into national data systems and functioning referral mechanisms, such innovations risk becoming short-lived, project-based interventions that bypass rather than strengthen services for displaced people.

**Table 1.**
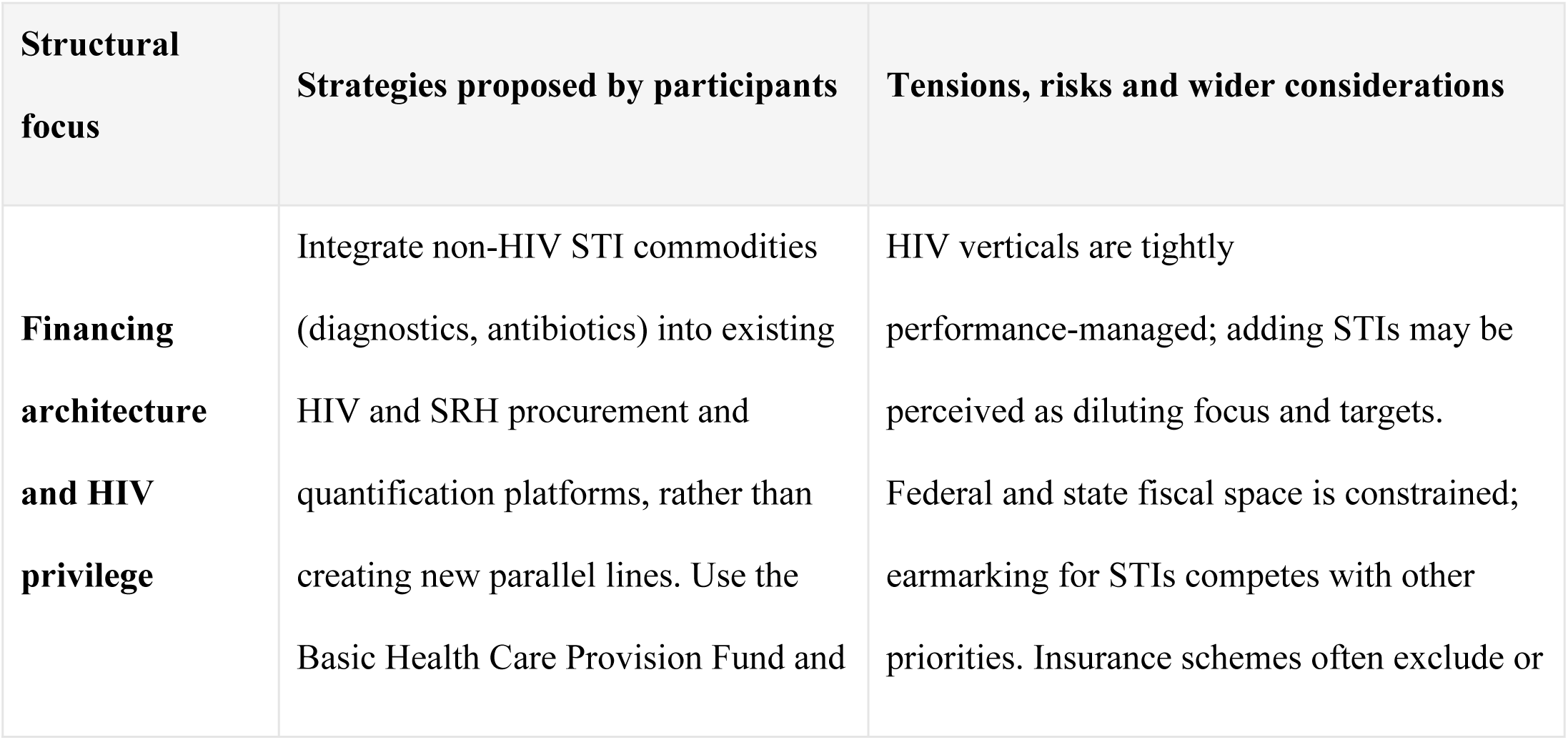
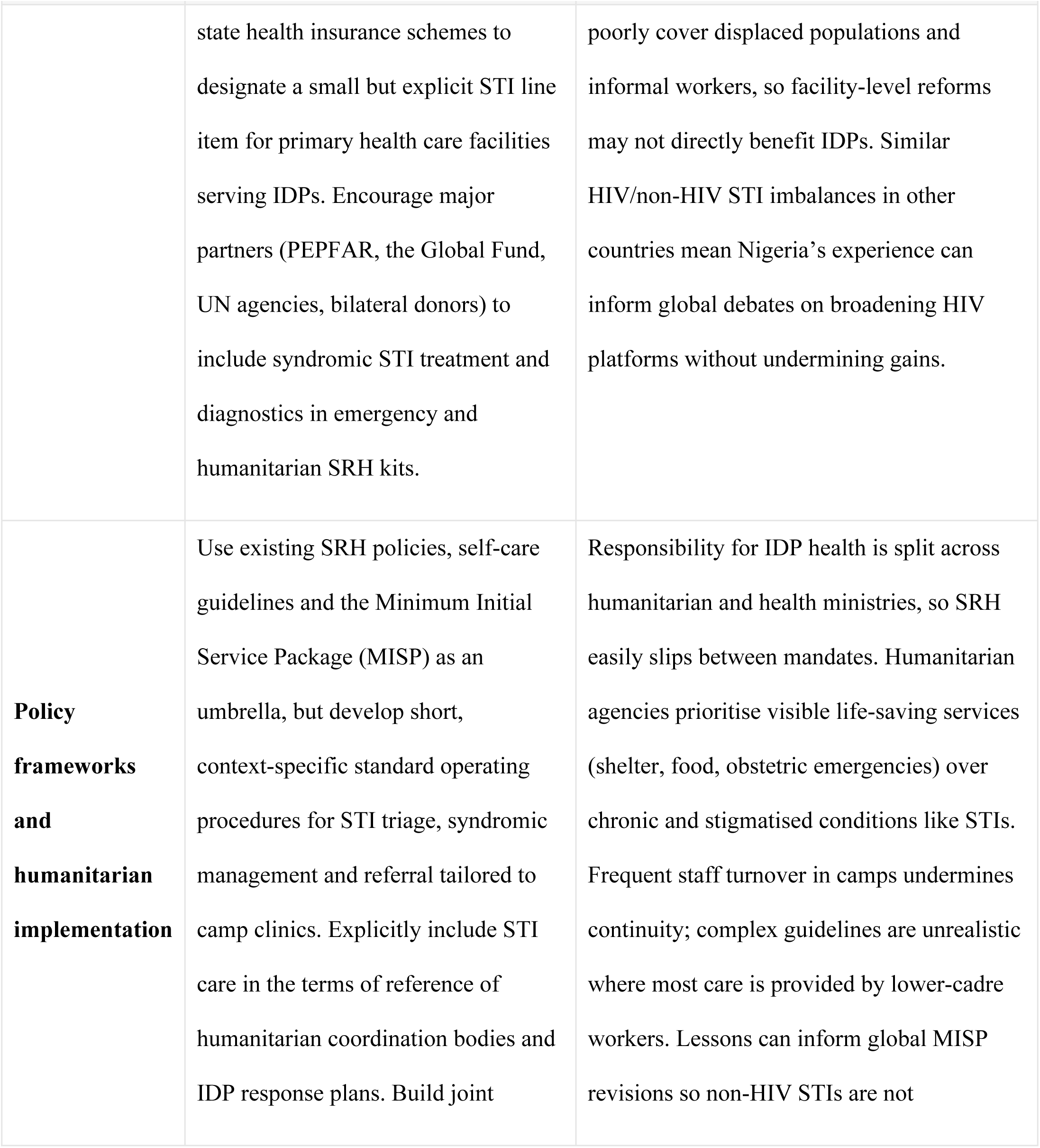
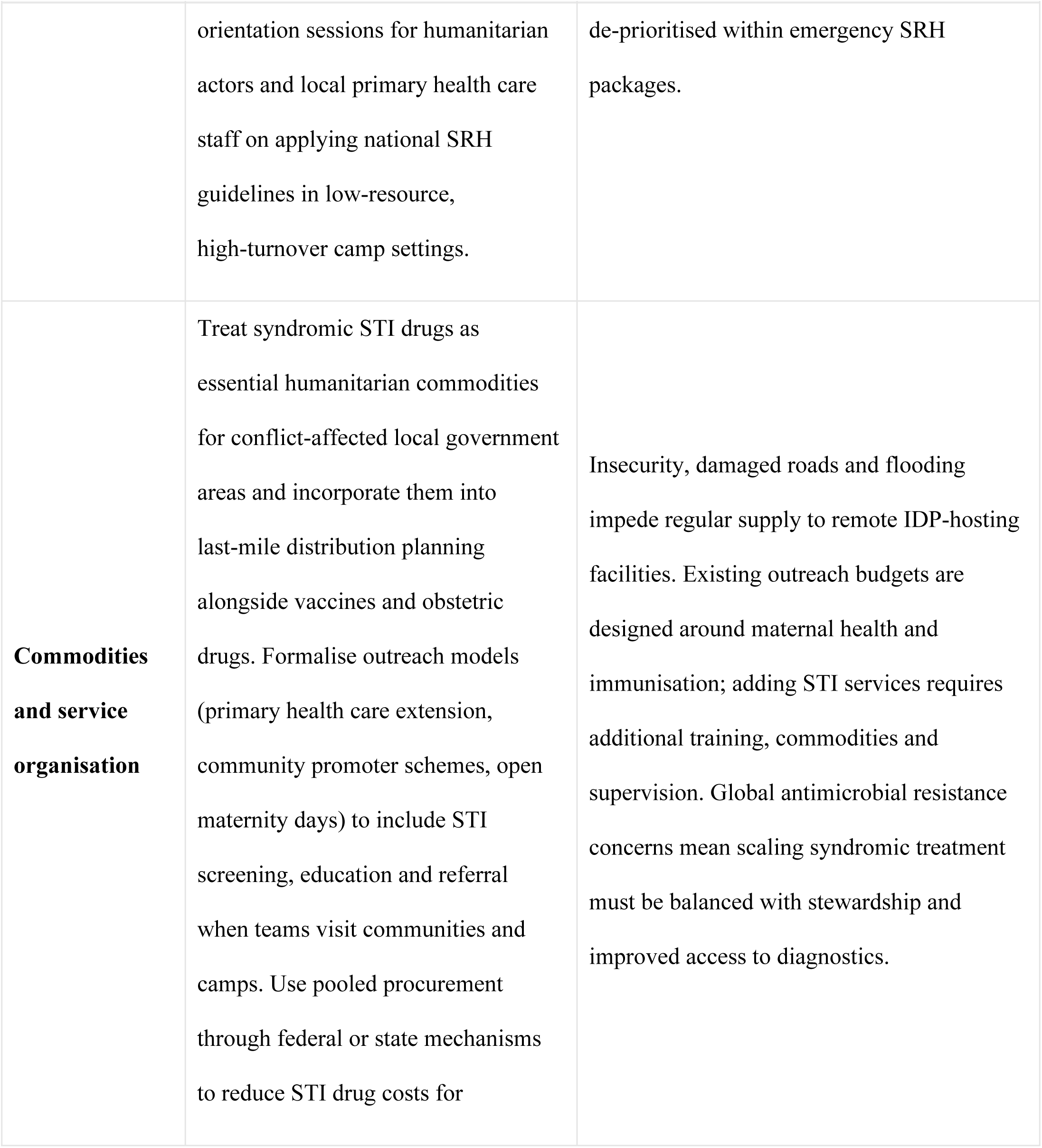
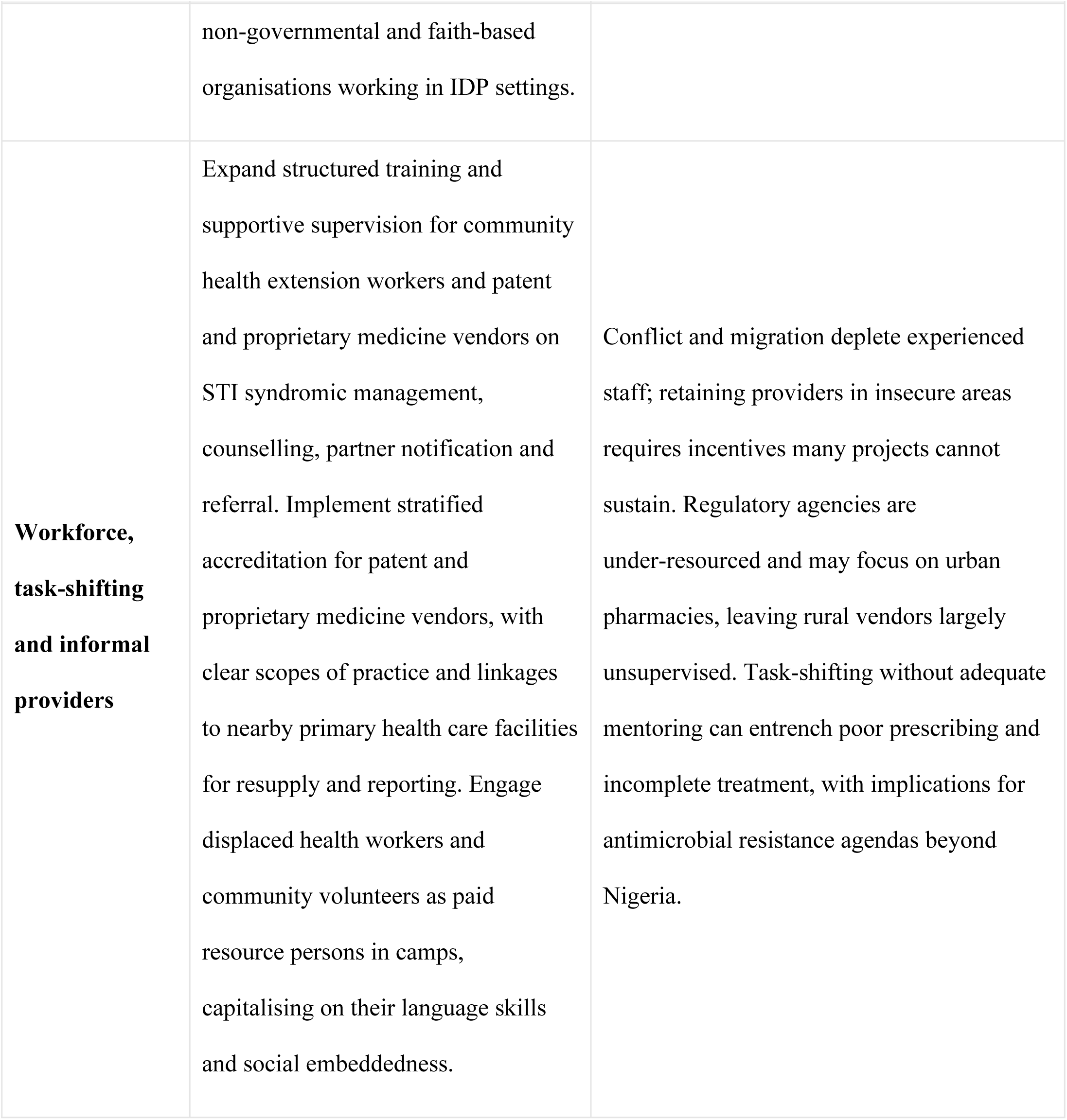
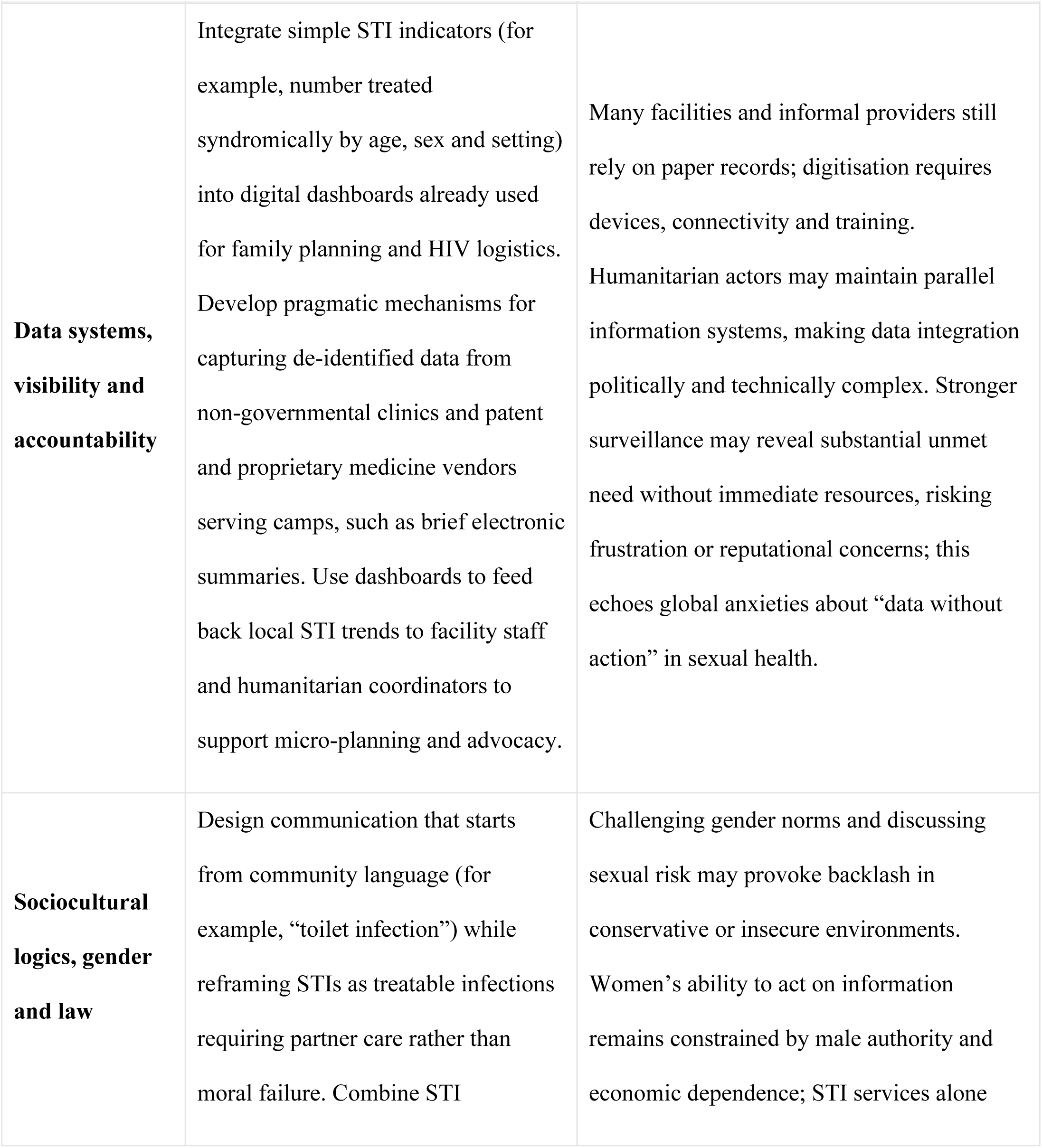
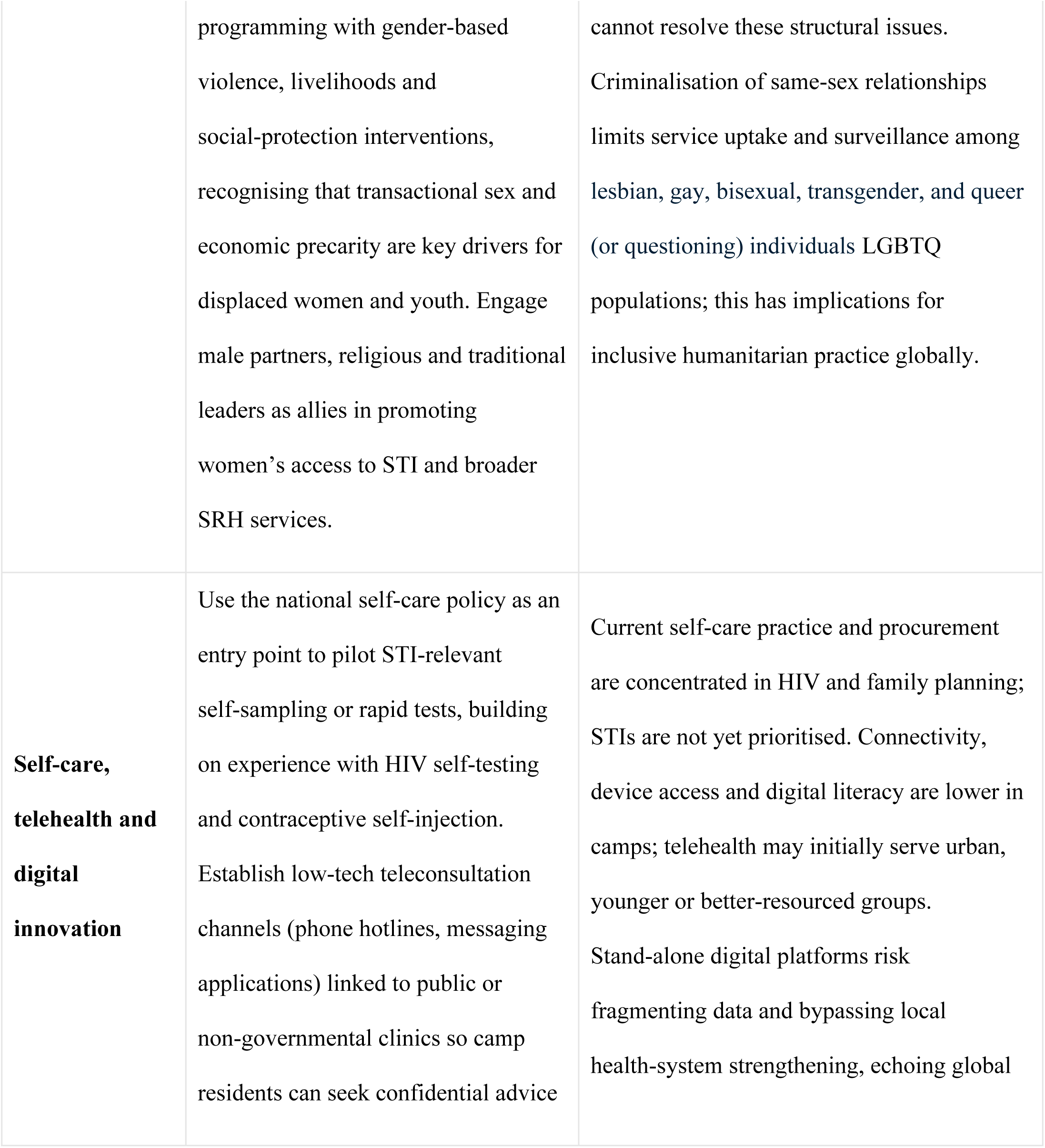
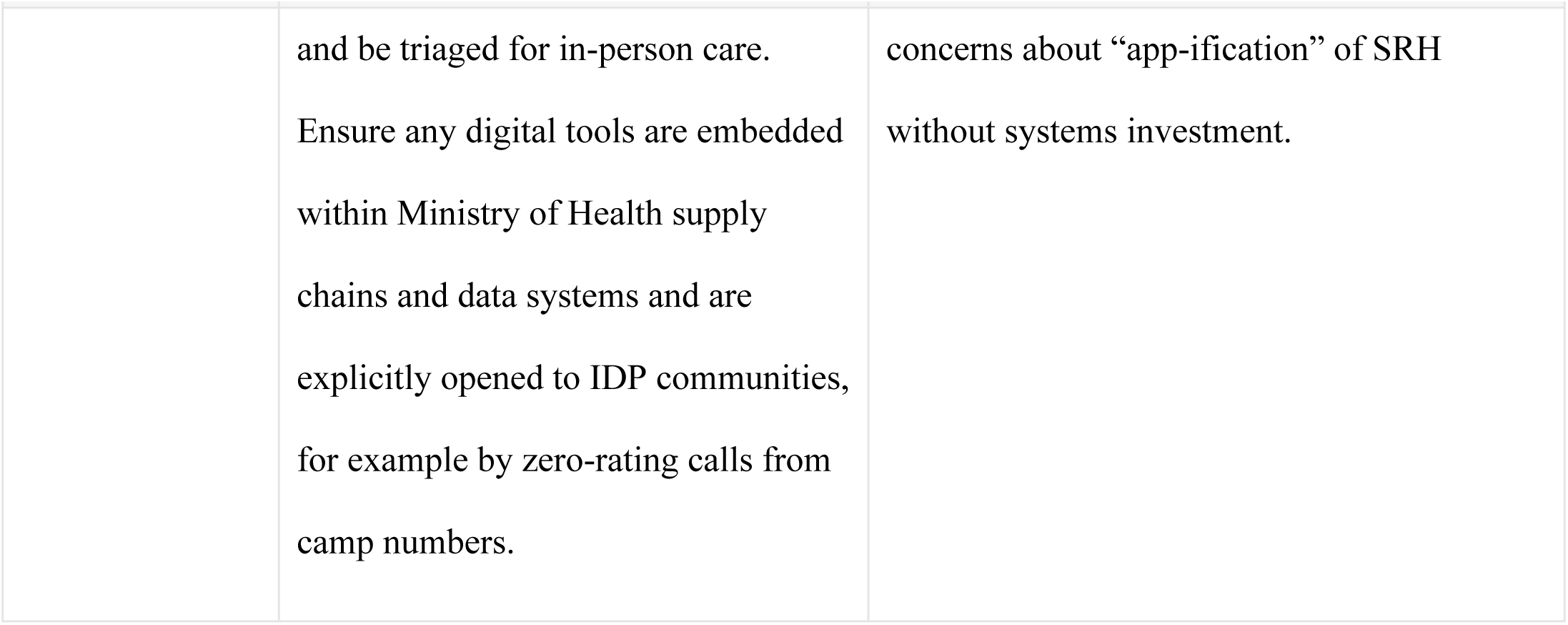
Structural Options for Integrating Non-HIV STI Services into Humanitarian Health Responses for IDPs in Nigeria.

| Structural focus | Strategies proposed by participants | Tensions, risks and wider considerations |
| --- | --- | --- |
| **Financing architecture and HIV privilege** | Integrate non-HIV STI commodities (diagnostics, antibiotics) into existing HIV and SRH procurement and quantification platforms, rather than creating new parallel lines. Use the Basic Health Care Provision Fund and | HIV verticals are tightly performance-managed; adding STIs may be perceived as diluting focus and targets. Federal and state fiscal space is constrained; earmarking for STIs competes with other priorities. Insurance schemes often exclude or |

| **Structural focus** | **Strategies proposed by participants** | **Tensions, risks and wider considerations** |
| --- | --- | --- |
|  | state health insurance schemes to designate a small but explicit STI line item for primary health care facilities serving IDPs. Encourage major partners (PEPFAR, the Global Fund, UN agencies, bilateral donors) to include syndromic STI treatment and diagnostics in emergency and humanitarian SRH kits. | poorly cover displaced populations and informal workers, so facility-level reforms may not directly benefit IDPs. Similar HIV/non-HIV STI imbalances in other countries mean Nigeria's experience can inform global debates on broadening HIV platforms without undermining gains. |
| **Policy frameworks and humanitarian implementation** | Use existing SRH policies, self-care guidelines and the Minimum Initial Service Package (MISP) as an umbrella, but develop short, context-specific standard operating procedures for STI triage, syndromic management and referral tailored to camp clinics. Explicitly include STI care in the terms of reference of humanitarian coordination bodies and IDP response plans. Build joint | Responsibility for IDP health is split across humanitarian and health ministries, so SRH easily slips between mandates. Humanitarian agencies prioritise visible life-saving services (shelter, food, obstetric emergencies) over chronic and stigmatised conditions like STIs. Frequent staff turnover in camps undermines continuity; complex guidelines are unrealistic where most care is provided by lower-cadre workers. Lessons can inform global MISP revisions so non-HIV STIs are not |
|  | orientation sessions for humanitarian actors and local primary health care staff on applying national SRH guidelines in low-resource, high-turnover camp settings. | de-prioritised within emergency SRH packages. |
| **Commodities and service organisation** | Treat syndromic STI drugs as essential humanitarian commodities for conflict-affected local government areas and incorporate them into last-mile distribution planning alongside vaccines and obstetric drugs. Formalise outreach models (primary health care extension, community promoter schemes, open maternity days) to include STI screening, education and referral when teams visit communities and camps. Use pooled procurement through federal or state mechanisms to reduce STI drug costs for | Insecurity, damaged roads and flooding impede regular supply to remote IDP-hosting facilities. Existing outreach budgets are designed around maternal health and immunisation; adding STI services requires additional training, commodities and supervision. Global antimicrobial resistance concerns mean scaling syndromic treatment must be balanced with stewardship and improved access to diagnostics. |
|  | non-governmental and faith-based organisations working in IDP settings. |  |
| **Workforce, task-shifting and informal providers** | Expand structured training and supportive supervision for community health extension workers and patent and proprietary medicine vendors on STI syndromic management, counselling, partner notification and referral. Implement stratified accreditation for patent and proprietary medicine vendors, with clear scopes of practice and linkages to nearby primary health care facilities for resupply and reporting. Engage displaced health workers and community volunteers as paid resource persons in camps, capitalising on their language skills and social embeddedness. | Conflict and migration deplete experienced staff; retaining providers in insecure areas requires incentives many projects cannot sustain. Regulatory agencies are under-resourced and may focus on urban pharmacies, leaving rural vendors largely unsupervised. Task-shifting without adequate mentoring can entrench poor prescribing and incomplete treatment, with implications for antimicrobial resistance agendas beyond Nigeria. |
| **Data systems, visibility and accountability** | Integrate simple STI indicators (for example, number treated syndromically by age, sex and setting) into digital dashboards already used for family planning and HIV logistics. Develop pragmatic mechanisms for capturing de-identified data from non-governmental clinics and patent and proprietary medicine vendors serving camps, such as brief electronic summaries. Use dashboards to feed back local STI trends to facility staff and humanitarian coordinators to support micro-planning and advocacy. | Many facilities and informal providers still rely on paper records; digitisation requires devices, connectivity and training. Humanitarian actors may maintain parallel information systems, making data integration politically and technically complex. Stronger surveillance may reveal substantial unmet need without immediate resources, risking frustration or reputational concerns; this echoes global anxieties about “data without action” in sexual health. |
| **Sociocultural logics, gender and law** | Design communication that starts from community language (for example, “toilet infection”) while reframing STIs as treatable infections requiring partner care rather than moral failure. Combine STI | Challenging gender norms and discussing sexual risk may provoke backlash in conservative or insecure environments. Women’s ability to act on information remains constrained by male authority and economic dependence; STI services alone |

| **Structural focus** | **Strategies proposed by participants** | **Tensions, risks and wider considerations** |
| --- | --- | --- |
|  | programming with gender-based violence, livelihoods and social-protection interventions, recognising that transactional sex and economic precarity are key drivers for displaced women and youth. Engage male partners, religious and traditional leaders as allies in promoting women's access to STI and broader SRH services. | cannot resolve these structural issues.<br>Criminalisation of same-sex relationships limits service uptake and surveillance among lesbian, gay, bisexual, transgender, and queer (or questioning) individuals LGBTQ populations; this has implications for inclusive humanitarian practice globally. |
| **Self-care, telehealth and digital innovation** | Use the national self-care policy as an entry point to pilot STI-relevant self-sampling or rapid tests, building on experience with HIV self-testing and contraceptive self-injection.<br>Establish low-tech teleconsultation channels (phone hotlines, messaging applications) linked to public or non-governmental clinics so camp residents can seek confidential advice | Current self-care practice and procurement are concentrated in HIV and family planning; STIs are not yet prioritised. Connectivity, device access and digital literacy are lower in camps; telehealth may initially serve urban, younger or better-resourced groups.<br>Stand-alone digital platforms risk fragmenting data and bypassing local health-system strengthening, echoing global |
|  | and be triaged for in-person care.<br>Ensure any digital tools are embedded within Ministry of Health supply chains and data systems and are explicitly opened to IDP communities, for example by zero-rating calls from camp numbers. | concerns about “app-ification” of SRH without systems investment. |

## Discussion

This study examined how structural, institutional, and sociocultural determinants shape STI service delivery for internally displaced persons (IDPs) in Nigeria. The findings suggest that inequities in STI care within displacement settings stem less from the absence of policy and more from structural asymmetries embedded in financing systems, commodity supply chains, workforce distribution, and social norms. Although Nigeria has made notable normative progress in sexual and reproductive health (SRH) and self-care policy development, implementation for non-HIV STIs remains constrained, particularly in humanitarian contexts.

### Funding Architecture and the Structural Privileging of HIV

Participants’ accounts of HIV having effectively overshadowed and marginalised the STI agenda reflect longstanding global health financing patterns in which vertically funded programmes shape national priorities and institutional authority [14,17]. Nigeria continues to carry one of the largest HIV burdens globally, with approximately 1.9 million people living with HIV and 1.69 million individuals receiving antiretroviral therapy (ART), and cascade indicators approaching the 95-95-95 targets [5]. This epidemiological context justifies sustained investment.

However, the structural privileging of HIV extends beyond disease burden. HIV programming benefits from vertically integrated procurement systems supported by PEPFAR, the Global Fund, and related bilateral initiatives. These mechanisms provide relatively stable supply chains, performance metrics, and largely free treatment at the point of care. In contrast, non-HIV STIs such as syphilis, gonorrhoea, and chlamydia lack equivalent commodity guarantees, national dashboards, or protected funding streams [1]. Empirical studies in Nigerian tertiary facilities indicate that patients treated for STIs frequently incur direct medication and consultation costs, with limited insurance coverage for syndromic management [18].

Participants’ observation that “for other STI generally they have to pay” therefore aligns with documented financing arrangements. Within Nigeria’s mixed public–private health system, non-HIV STI care is predominantly financed through out-of-pocket expenditure. This divergence mirrors broader global evidence showing that vertical HIV programming can coexist alongside comparatively underfunded STI services [16].

In displacement contexts, these structural imbalances are amplified. With over three million internally displaced persons nationwide [6], humanitarian programming often prioritises HIV, maternal health, and emergency obstetric care. Broader STI programming remains peripheral. Funding architecture thus operates as a determinant of visibility: infections supported by dedicated financing are counted, monitored, and subsidised, while others remain fragmented and less visible within both national and humanitarian systems. Recent contractions in HIV and broader development assistance for health following the COVID-19 pandemic further complicate this landscape, with declining global HIV budgets and constrained fiscal space in many low- and middle-income countries. These trends suggest that the financing asymmetries described by participants are unlikely to narrow without deliberate efforts to integrate non-HIV STI services into existing funding platforms.

The January 2025 United States foreign assistance freeze, which suspended and in several cases terminated PEPFAR-funded programme activities in Nigeria, offers a stark empirical illustration of the systemic fragility our participants described [10]. Implementing partners supporting antiretroviral therapy, key population services, and One-Stop Shops received stop-work orders with no transition arrangements in place, creating immediate supply chain disruptions [10,11]. For internally displaced populations — who already lacked alternative access to subsidised STI care — this disruption compounds an existing structural deficit rather than creating a new one. Critically, it also demonstrates that the risk embedded in vertically financed, donor-dependent programming is not hypothetical: our findings document the conditions that made Nigeria’s HIV-anchored sexual health architecture so exposed. A system that had already marginalised non-HIV STI services within displacement settings now faces the simultaneous destabilisation of its dominant financing pillar. This underscores the argument, derived from participant accounts and supported by the structural determinants framework [13], that financing diversification and domestic integration of STI services are not aspirational goals but urgent system priorities.

### Policy Proliferation and Implementation Gaps in Displacement Settings

Nigeria’s SRH policy landscape is extensive. National guidelines on self-care [9], task-shifting [15], and commitments aligned with the Minimum Initial Service Package [7] formally recognise vulnerable populations, including IDPs. WHO guidance similarly promotes HIV self-testing and mpling as strategies to expand access [3].

Despite these frameworks, participants consistently described a gap between policy articulation and operational delivery. Implementation in IDP camps is constrained by weak laboratory infrastructure, unstable supply chains, limited regulatory oversight of informal providers, and persistent workforce shortages. These findings align with evidence that humanitarian health systems often prioritise rapid deployment and emergency stabilisation over long-term integration and diagnostic capacity, leading to reliance on syndromic management rather than laboratory-confirmed diagnosis [1].

HPV self-sampling and STI self-collection have demonstrated high acceptability in several sub-Saharan African contexts. A community-based study in the West Region of Cameroon reported strong acceptance of home-based HPV self-sampling, while a rural Delta State study in Nigeria documented an acceptability rate of approximately 93% and high confidence in correct device use [24,25].

### Health System Constraints: Commodities, Workforce, and Data

Three interlocking constraints emerged as central to participants’ accounts: commodity insecurity, workforce shortages, and fragmented data systems.

### Commodity Insecurity

While antiretroviral therapy and family planning commodities benefit from protected procurement mechanisms, no national free-drug scheme exists for syndromic STI treatment outside HIV programming [1]. In displacement settings, this often results in reliance on PPMVs and private pharmacies. Evidence from sub-Saharan Africa indicates that STI-related care can generate significant out-of-pocket expenditure, including catastrophic spending among vulnerable populations [23]. In Nigeria, limited insurance coverage among informal and displaced populations further entrenches these inequities.

### Workforce Shortages and Task-Shifting

Conflict-affected states experience intensified workforce attrition due to insecurity and migration. National strategies endorse task-shifting to community health extension workers and PPMVs as a means of maintaining service coverage [19,15,20]. While this approach expands geographic reach, concerns persist regarding supervision, quality assurance, and antimicrobial stewardship [2,21]. In fragile settings, these regulatory gaps may compound risks of incomplete treatment and antimicrobial resistance.

### Data Invisibility

Participants’ emphasis on data gaps reflects broader global concerns regarding surveillance for non-HIV STIs [22]. Nigeria’s HIV burden has been periodically refined through national surveys such as NAIIS [4]. No comparable population-based surveillance system exists for the full STI spectrum. Routine information systems do not consistently capture STI episodes managed in private or humanitarian channels.

This statistical invisibility reinforces policy neglect. Conditions that are not routinely measured remain difficult to prioritise. Participants’ recommendation to integrate STI indicators into digital dashboards aligns with growing international emphasis on strengthening digital SRH data systems [1]. Participants’ reliance on syndromic “triple therapy” in the absence of culture capacity mirrors global concerns that empiric treatment without diagnostics may contribute to antimicrobial resistance, particularly for gonorrhoea, in settings where surveillance remains weak.

### Sociocultural Logics, Gender, and Structural Vulnerability

Narratives linking STI symptoms to “toilet infection” illustrate how local explanatory models shape health-seeking behaviour. While sanitation-related attributions may reduce sexual stigma, epidemiological evidence indicates that sexual transmission remains the primary driver of most STIs [26]. Misattribution may delay partner notification and appropriate treatment.

Gender norms further shape access. In parts of northern Nigeria, women’s healthcare decisions may require male consent [27]. Within displacement settings, these norms intersect with economic precarity and heightened exposure to gender-based violence [28]. Participants’ accounts of transactional sex and untreated infection underscore how structural vulnerability translates into long-term reproductive consequences.

Legal constraints also shape access. Criminalisation of same-sex relationships restricts service utilisation and surveillance among LGBTQ populations [2]. In humanitarian contexts structured around heteronormative family models, such populations may remain both socially and statistically marginalised.

Taken together, these dynamics support a structural determinants interpretation [13]. Funding asymmetries, gender hierarchies, legal exclusion, and displacement intersect to shape who is exposed, who seeks care, and whose infections are formally recognised.

### Digital Futures and Equity Considerations

Participants anticipated digital platforms and telehealth as emerging avenues for expanding access. The post-COVID expansion of teleconsultation and app-supported HIV self-testing confirms the growing role of digital health [1]. Youth-focused integrated testing interventions have demonstrated promising uptake [26].

However, digital access is uneven. Connectivity, device ownership, and digital literacy remain lower in displacement settings [29,30]. Without integration into public insurance mechanisms and humanitarian benefit packages, telehealth may reinforce rather than reduce inequities.

Digital innovation should therefore complement strengthened primary healthcare systems rather than substitute for them. Integration into Federal Ministry of Health dashboards and alignment with national supply chains would be necessary to avoid creating parallel, project-based data systems. The rapid scale-up of teleconsultation and app-supported HIV self-testing during the COVID-19 pandemic shows that digital SRH adaptations can be implemented quickly when there is political will and donor support. Yet participants’ accounts of limited connectivity, device access and digital literacy in camps underscore that such innovations may bypass displaced populations unless explicitly integrated into public financing and humanitarian benefit packages.

### Strengths and Limitations

This study provides institutional insight into STI service delivery within Nigeria’s displacement context, drawing on perspectives from national, sub-national, and humanitarian actors. By focusing on structural determinants, it highlights dynamics that may not be visible through epidemiological data alone.

However, the study reflects the perspectives of policymakers and programme actors rather than IDPs themselves. Future research should incorporate the lived experiences of displaced populations to deepen understanding of service access barriers. In addition, the policy environment has continued to evolve since data collection, and implementation conditions may have shifted in response to changing funding and geopolitical dynamics.

### Conclusion

STI care for internally displaced persons in Nigeria is shaped less by policy absence than by structural determinants embedded in financing architectures, commodity systems, workforce distribution, data infrastructures and sociocultural norms. While HIV’s programmatic dominance is epidemiologically grounded, our findings suggest that it has inadvertently contributed to the marginalisation of non-HIV STIs within humanitarian programming. Nigeria’s policy frameworks on self-care, task-shifting and digital innovation provide an important normative foundation; yet without sustained commodity security, strengthened surveillance beyond HIV, regulatory oversight of informal providers and more equitable financing mechanisms, these frameworks are unlikely to achieve substantive impact in displacement settings.

Advancing equitable STI care for displaced populations will require deliberate integration of comprehensive STI services into humanitarian and national financing streams, stronger and more inclusive surveillance systems, and programming that addresses gendered and other structural vulnerabilities. These imperatives have become more urgent, not less, since data collection. The 2025 PEPFAR disruption has exposed in real time the fragility of a sexual health architecture built around a single vertical financing pillar [10,11]; the WHO’s 2024–2025 progress data confirm that global syphilis cases are rising, STI surveillance capacity remains weak across the African region, and the 2030 STI targets are off-track [12]. Nigeria’s internally displaced populations sit at the intersection of all these pressures. By situating STI service delivery within its broader structural context, this study identifies internally displaced populations as a critical frontier for equitable sexual health policy in Nigeria, while acknowledging that our analysis—based on key informant perspectives—should be complemented by future work incorporating displaced people’s own experiences of STI care.

## Data Availability

The qualitative interview data underlying this article are not publicly available because they contain potentially identifiable information from a small group of experts and officials discussing health systems, policies, and service delivery in Nigeria. Participants were recruited in their professional roles (e.g. senior government, UN and NGO positions), and even after pseudonymisation it would be possible to re?identify individuals and organisations based on contextual details. Public data sharing would therefore contravene the assurances of confidentiality provided in the informed consent process and the conditions under which ethics approval was granted. De?identified excerpts relevant to the analysis are included in the article additional anonymised segments may be shared on reasonable request to the corresponding author, subject to institutional ethics approval and, where necessary, renewed consent from participants.

## Acknowledgments

I am deeply grateful to the many stakeholders who generously shared their time and insights for this study, including representatives from federal and state ministries, international and local non-governmental organisations, United Nations agencies, professional associations, and clinicians providing care to internally displaced populations in Nigeria.

